# Pandemic-Potential Viruses are a Blind Spot for Frontier Open-Source LLMs

**DOI:** 10.64898/2025.12.04.25341642

**Authors:** Laura Luebbert, Yasha Ektefaie, Arya S. Rao, Colby Wilkason, Dolo Nosamiefan, Olivia Achonduh-Atijegbe, Harouna Soumare, Adefoye Precious Adebayo, Olufemi Olulaja, Judith Amadi, Nicholas Oyejide, Funmilayo Olayiwola, Etim Henshaw, Yusuf Okocha, Nkechinyere Nwachukwu, Elechi Friday Ewah, Sylvanus Okoro, Ebenezer Nwakpakpa, Peter Okokhere, Kelly Iraoyah, Joseph Okoeguale, Ireti Dada, Andy Burris, Karlie Zhao, Ellory Laning, Chase van Amburg, Paul Cronan, Ben Fry, Christian Happi, Al Ozonoff, Pardis C. Sabeti

## Abstract

We study large language models (LLMs) for front-line, pre-diagnostic infectious-disease triage, a critically understudied stage in clinical interventions, public health, and biothreat containment. We focus specifically on the operational decision of classifying symptomatic cases as *viral* vs. *non-viral* at first clinical contact, a critical decision point for resource allocation, quarantine strategy, and antibiotic use. We create a benchmark dataset of first-encounter cases in collaboration with multiple healthcare clinics in Nigeria, capturing high-risk viral presentations in low-resource settings with limited data. Our evaluations across frontier open-source LLMs reveal that (1) LLMs underperform standard tabular models and (2) case summaries and Retrieval Augmented Generation yield only modest gains, suggesting that naïve information enrichment is insufficient in this setting. To address this, we demonstrate how models aligned with Group Relative Policy Optimization and a triage-oriented reward consistently improve baseline performance. Our results highlight persistent failure modes of general-purpose LLMs in pre-diagnostic triage and demonstrate how targeted reward-based alignment can help close this gap.

## 1 Introduction

Outbreak control often hinges on the uncertain hours before test results arrive, when isolation and resource allocation decisions cannot wait. In this pre-diagnostic window, clinicians must act on partial and uncertain information, making decisions about whether to isolate the patient, escalate testing, and allocate limited resources. A simple but critical distinction is whether an illness is likely *viral* or *non-viral*. That signal can shape whether a patient is prioritized for scarce PCR assays or sequencing, guide use of antibiotics, and determine the level of protective measures needed to prevent onward transmission.

Despite its importance, this decision space is almost entirely absent from existing AI benchmarks. Gold-standard clinical datasets such as MIMIC-IV [1] reflect retrospective, high-income tertiary care populations where diagnosis has already been established. Outbreak-oriented resources such as GISAID [2] or WHO [3] case counts capture pathogen dynamics at the population level, but lack patient-level data at presentation. There is little open evidence on how machine learning models perform when confronted with the actual conditions of frontline triage: heterogeneous patients, incomplete measurements, and uncertainty about the causative pathogen.

To establish a baseline for performance on these tasks, we created an initial dataset collected through our initiative Sentinel, a global health organization addressing emerging infectious diseases in Africa. By partnering with locally led healthcare clinics in Nigeria, we collected structured clinical features available at or near presentation: vital signs, point-of-care rapid tests, early laboratory values, epidemiologic exposures (travel, contact history, occupational risks), demographics, and comorbidities. Each encounter is linked to definitive outcomes when testing is performed, producing a realistic view of what clinicians know (and what they do not) at the time critical decisions are made.

We study LLMs in a setting largely absent from prior benchmarks: front-line, pre-diagnostic infectious-disease triage (Figure 1). We address this gap with the following contributions:

1. We **introduce a dataset of 13,628 first-encounter cases** from locally led networks across multiple West African sites, with presentation-time clinical features and confirmatory outcomes.
2. We establish an open, reproducible benchmark for the operational decision *viral* vs. *non-viral*, evaluating state-of-the-art (SOTA) open-source LLMs under four prompting regimes (structured JSON; medical case summaries; feature-text prompting; patient-RAG) against a tabular baseline (random forest). We find that **all tested large language models underperform standard statistical baselines**.
3. To close this gap, we align Gemma-4B with Group Relative Policy Optimization (GRPO) using a triage-specific reward [4]. **Our aligned model improves baseline performance and highlights the promise of reward-based alignment in pre-diagnostic triage**.

**Figure 1:**
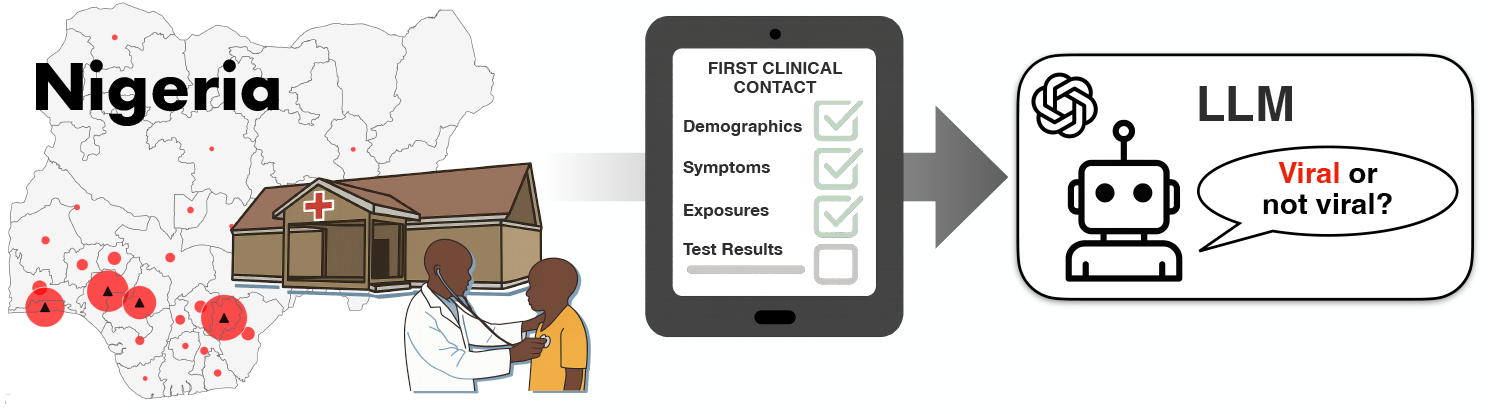
Schematic overview of the study design. We collect first contact data from a broad range of patients in Nigeria, and evaluate large language models on their ability to classify symptoms as *viral* or *non-viral*. Despite representing a critical decision boundary for clinicians, LLMs struggle to surpass traditional baselines.

We prioritize open-source models to enhance reproducibility, safeguard data privacy, and ensure applicability in low-resource settings. We also release our evaluation harness, and the dataset will be made publicly available after de-identification and governance review by Sentinel.

## 2 Related Work

### LLMs in Medicine

LLMs have spurred great interest in clinical medicine on tasks including clinical knowledge retrieval, diagnosis, management, and more. Off-the-shelf LLMs have achieved impressive results on standardized medical exams and other question-answering tasks [5]. Tailored models have also achieved impressive results: MedPaLM 2 [6] reports near-clinician accuracy on standardized medical exams. Community efforts such as ChatDoctor [7] and MedAlpaca [8] explore instruction tuning of general-purpose LLMs on curated medical corpora. BioMedLM [9] extends this line by pretraining directly on biomedical literature, while TxAgent [10] introduces a tool-augmented agent framework that integrates external biomedical knowledge into LLM reasoning. Despite these advances, most evaluations focus on exam-style or post-diagnostic settings; none target the high-stakes, first-encounter infectious-disease triage scenario we study.

### Benchmarks for LLMs in Medicine

Most benchmarks for clinical LLMs emphasize static knowledge recall rather than decision making under uncertainty at presentation. Exam-derived datasets such as MedQA (USMLE) [11], MedMCQA [12], and the medical slice of MMLU [13] primarily assess factual recall and diagnostic reasoning. PubMedQA [14] targets literature-grounded Q&A, and USMLE Self-Assessment [15] adds standardized items. To move beyond single-turn exams, Rao *et al*. [16] introduced multi-turn clinical encounters that mimic full clinical workflows using vignette-derived cases. Other resources broaden modalities: MedXpertQA [17] supplies expert-curated case questions, and VQA-RAD [18] benchmarks visual Q&A over radiology images. Parallel to these, the PhysioNet ecosystem [19], including MIMIC/eICU cohorts and large waveform banks as well as the annual PhysioNet Challenges, has enabled rigorous evaluation on supervised, time-series prediction tasks, e.g., arrhythmia classification ans sepsis/mortality forecasting. However, these datasets are largely retrospective, inpatient, and post-diagnostic, and most LLM evaluations on them do not reflect first-contact decision constraints. Most recently, HealthBench [20] advances rubric-scored, multi-turn clinician–model conversations across specialties and countries, but remains primarily synthetic and disconnected from real-world first-encounter data. A dedicated benchmark for front-line, *pre-diagnostic* infectious-disease triage has been lacking. Our dataset is designed to fill this gap.

### Reinforcement Learning for Medical LLMs

Reinforcement learning (RL) has emerged as a strategy to improve LLM performance beyond likelihood-based finetuning by directly optimizing medical-specific preferences and safety criteria. In the medical domain, MedRLVR [21], MedU1 [22], and Rubric-as-Reward [23] show that reward-driven optimization can improve alignment and down-stream performance even with limited supervision. Building on this direction, we apply GRPO to align Gemma-4B to front-line infectious-disease triage signals, and demonstrate consistent gains [4].

## 3 Methods

### 3.1 Dataset

The dataset used to evaluate whether models could predict *viral* vs. *non-viral* illness was collected by our initiative, Sentinel, a pathogen surveillance and technology program in West Africa run by the Broad Institute of MIT and Harvard and the Institute of Genomics and Global Health (IGH) at Redeemer’s University in Nigeria. Clinical data were collected across four partner hospitals in Nigeria (Figure 2D). The dataset comprises 13,628 patient encounters, providing a large and diverse cohort spanning multiple Nigerian states, with most patients residing in Ebonyi, Ondo, and Lagos States (Figure 2A and D). The average patient population was relatively young, with 46.3% aged 25–50 years and 34.7% under 25 years, and included more women (67.9%) than men (32.0%). Patients were recruited primarily through outpatient and emergency departments, particularly the General Outpatient Department (GOPD) and Accident and Emergency (A&E) units.

**Figure 2:**
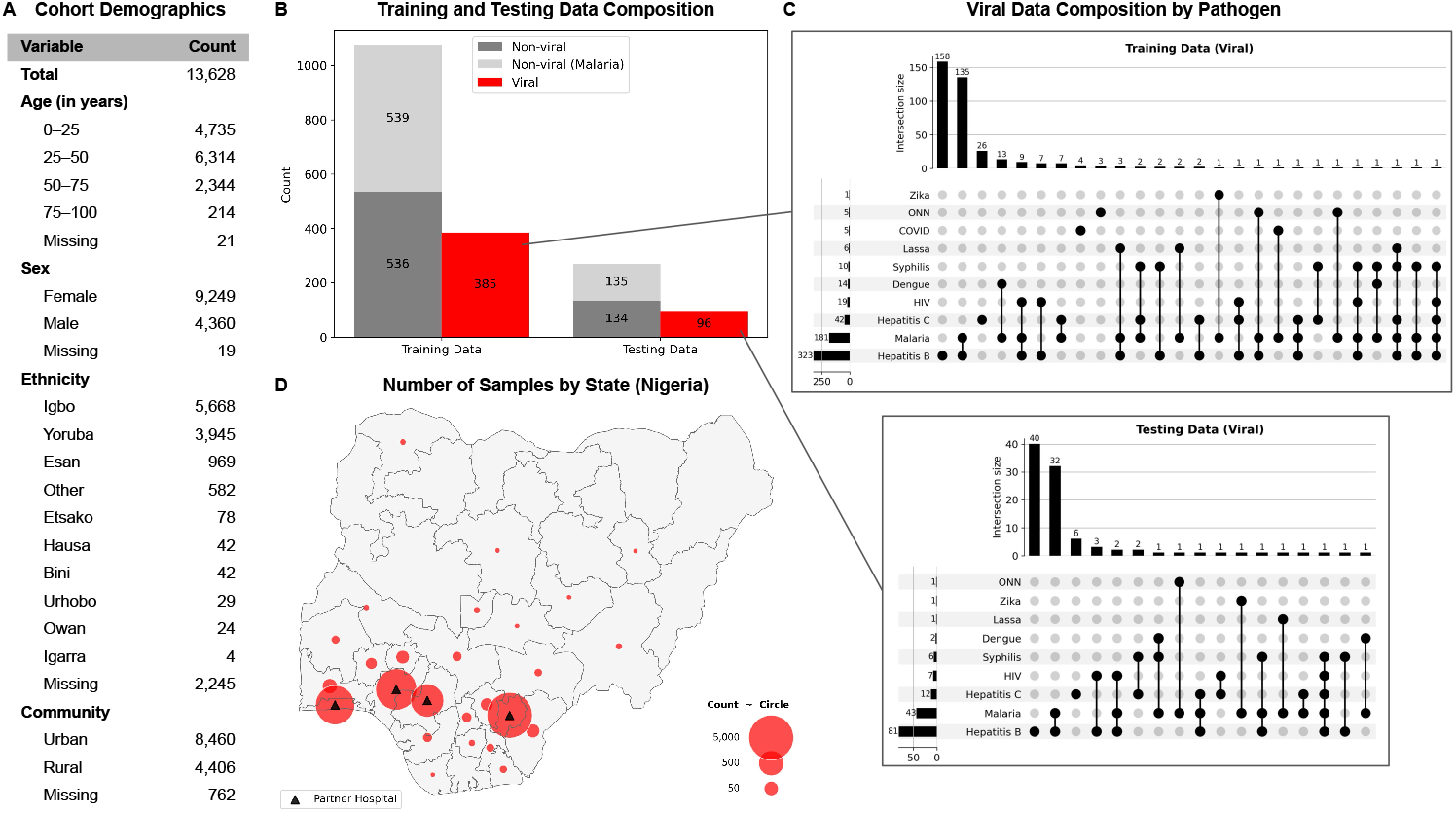
Cohort demographics and training and testing data composition. **(A)** Age, sex, ethnicity, and community distribution for the 13,628 patients enrolled. **(B)** Training and testing data composition to evaluate *viral* vs. *non-viral* predictions. *Viral* samples were defined as those with at least one confirmed viral diagnosis (excluding HIV monoinfection), inclusive of co-infections (n=481). *Non-viral* samples were defined as (i) all patients with ≥ 5 confirmed negative viral tests and no positive test (n=674), and (ii) 674 randomly selected malaria-positive patients without confirmed co-infections. Malaria-positive patients were included to evaluate the clinically important distinction between viral infection and malaria. **(C)** Confirmed viral diagnoses in training and testing sets by pathogen. **(D)** Geographic distribution of patients across Nigeria.

Beyond demographics, the dataset captures a rich set of clinical, epidemiological, and social features. Structured variables include vital signs (81% of patients presented with fever symptoms, with 17.8% exhibited a measured temperature ≥ 38 °C), comorbidities (hypertension, diabetes, tuberculosis, among others), and detailed symptom profiles spanning neurological, respiratory, and gastrointestinal systems. Exposure data document housing conditions, water access, animal contacts, and healthcare-related risks, reflecting important social and environmental determinants of infection risk. A detailed breakdown of all clinical variables is archived on Zenodo (DOI: 10.5281/zenodo.17065617).

To establish definitive viral diagnoses, suspected cases underwent laboratory confirmation using rapid diagnostic tests (HIV, Malaria, Hepatitis B, Hepatitis C, Syphilis, COVID), PCR assays (Yellow fever, Lassa, Ebola, Marburg, Zika, West Nile, Crimean-Congo Hemorrhagic Fever, Rift Valley, Dengue, O’nyong’nyong virus (ONN), COVID, Mpox), and/or blood film microscopy (Malaria) (Figure 2B and C).

Taken together, this dataset provides a comprehensive snapshot of patient presentation at the point of care, capturing the information clinicians have when making triage and diagnostic decisions. Its structured tabular format allows direct comparison of predictive models across a wide range of features, while linkage to definitive diagnostic outcomes creates a realistic benchmark for evaluating model performance. This resource therefore enables rigorous testing of whether models can predict *viral* vs. *non-viral* infection at first clinical presentation.

### 3.2 Data preprocessing

To reduce the influence of implausible outliers, we applied a series of common-sense filters to the clinical metadata (see Appendix A.1). After applying these filters, a total of 13,628 patient records were kept for downstream analysis. To prevent information leakage, we removed variables that directly encoded pathogen identity or otherwise provided the models with trivial shortcuts to the correct label. In particular, any feature that explicitly revealed the diagnostic outcome was excluded. For each pathogen, the results of multiple diagnostic assays were consolidated into a single binary indicator (positive/negative).

#### Ground-truth labels and training/testing splits

*Viral* samples were defined as those with at least one confirmed viral diagnosis (excluding HIV monoinfections to focus on acute rather than chronic infections), including co-infections (n=481). *Non-viral* samples were defined as (i) all patients with ≥ 5 confirmed negative viral tests and no positive test (n=674), and (ii) 674 randomly selected malaria-positive patients without confirmed co-infections. Malaria-positive patients were included to evaluate the clinically important distinction between viral infection and malaria. The data was then split into training (80%) and testing (20%) sets using a stratified train–test split (see Appendix A.1 for additional information).

### 3.3 Random forest and XGBoost

To establish a baseline, we trained ensemble classifiers (Random Forest and XGBoost) to distinguish viral from non-viral illness using structured clinical and epidemiological features. All experiments were conducted in Python 3.12 using *scikit-learn* (v1.7.1) [24] and *xgboost* (v2.1.1) [25]. Additional details on the encoding of variables and model training are provided in Appendix A.1.

Model performance was assessed on the held-out testing dataset. Probabilistic outputs (predict_proba) were used to compute Receiver Operating Characteristic (ROC) curves and the area under the ROC curve (AUC). The optimal classification threshold for each model was determined using Youden’s *J* (*J* = sensitivity (− 1 − specificity)). Sensitivity, specificity, and accuracy were then calculated at this threshold.

### 3.4 Baseline LLM inference

LLMs were deployed locally on multi-GPU compute servers (NVIDIA RTX A6000). Inference was performed through a standardized local API interface using the Ollama framework (v0.11.4), with a generation temperature of 0.5, top-*k* sampling set to (40), and default reasoning effort parameters. These settings were chosen to balance stability, diversity, and interpretability across model runs.

#### Prompting strategies

The system prompt used for all LLMs is shown in Box A.1 (Appendix). In the prompt, `field_definitions` is a structured JSON, in the format provided by REDCap (Research Electronic Data Capture), which provides additional information about the clinical and epidemiological features, such as:

~~~
{“Field”:”unit_crf”,”Choices”:”1 GOPD|2 Pediatrics|3 A&E|4 Other”},
{“Field”:”age_crf”,”Note”:”Years; 0=0–11 mo, 1=12–23 mo, etc.”},
{“Field”:”sex_crf”,”Choices”:”1 Male|2 Female”}, …
~~~

The user prompts were constructed as shown in Table A1 (Appendix) for each context setting, where `patient_text` contains the current patient data in structured JSON format, for example:

~~~
{“age_crf”: 25, “sex_crf”: 2, “unit_crf”: 1, …}
~~~

To rule out the possibility that model performance was limited by difficulties in interpreting structured JSON, we also evaluated the LLMs using inputs (`patient_text` and `field_definitions`) converted to natural language. Using the example above, the data would be reformatted as:

~~~
“The patient is 25 years old. The patient is female. The patients hospital unit/department is A&E…”
~~~

This modification did not affect the performance of the model (Figure S1).

#### Medical Context Summaries

For the medical context setting, we built a distilled `knowledge_summary` from labeled training data using a two-stage summarization pipeline designed to ensure all information fits within the model’s context window (see Appendix A.1 for additional information).

#### Retrieval Augmented Generation

We built a retrieval-augmented generation (RAG) vector store directly from tabular data. Each row of the dataset was converted into a compact JSON string, capturing selected columns as a structured record. To represent the records in vector space, we embedded the JSON strings using the Sentence Transformers library (v5.1.0) with the `all-MiniLM-L6-v2` model. Embeddings were generated in batches of up to 128 examples, and by default were normalized to unit length so that L2 distances in the index correspond to cosine similarity. We recorded the model name and normalization settings to ensure reproducibility. We then built a FAISS index using *faiss-gpu* (v1.11.0) [26] and an `IndexFlatL2` backend. With normalized embeddings, this index provides cosine-equivalent retrieval while keeping the configuration simple and parameter-free. Results are returned as text–metadata–score triplets, where scores reflect L2 distances.

At inference, the current patient’s data was encoded into the same embedding space and queried against the index to retrieve the top k (here, 20) most similar prior cases. Rather than supplying full neighbor records to the model, we extracted only their binary viral diagnosis labels (1 = positive, 0 = negative), yielding a compact list of labels representing the empirical distribution of outcomes among the nearest neighbors. This retrieved block of labels was inserted into the LLM prompt alongside instructions for its interpretation (Table A1), and the current patient’s data.

To evaluate RAG alone, we retrieved the 20 most similar cases from the FAISS index and computed the weighted average of their viral diagnosis labels. Predictions were then assigned as positive if the weighted average exceeded 0.5 and negative otherwise. This provided a direct baseline for the contribution of retrieval-based similarity independent of the language model.

### 3.5 GRPO Finetuning of Gemma-4B

We fine-tuned the `Gemma-3-4B-IT` model [27] using Group Relative Policy Optimization (GRPO) [4]. To reduce compute overhead, we applied LoRA adapters [28] while keeping the base model frozen, and used a parallel frozen reference model to impose KL regularization. Each training example consisted of a structured patient record rendered as a synthetic vignette within a chat template. For each prompt, the policy generated four candidate responses via nucleus sampling. Responses were scored with a domain-specific reward function. GRPO then optimized the policy by combining the PPO objective with the structured reward signal and a KL penalty to maintain stability [4]. This setup aligns the model’s free-form generations with clinically meaningful diagnostic reasoning while remaining efficient to train at scale.

#### Reward

We define a reward function *R* ∈ [0, 1] that evaluates the model’s predicted probability of viral status. Let *p* ∈ (0, 1) denote the predicted probability of the case being viral, and *y* ∈ {0, 1} the ground-truth label. We compute a signed logit margin

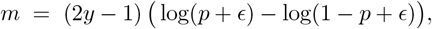

where *ϵ* is a small constant for numerical stability. The margin is positive when the model assigns higher probability to the correct class, and its magnitude increases as predictions move further from 0.5. The final reward is obtained by mapping the margin into the unit interval via a scaled hyperbolic tangent:

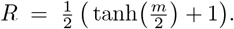

This formulation ensures bounded, smooth rewards: predictions near chance (*p* ≈ 0.5) yield rewards close to 0.5, confidently correct predictions approach 1, and confidently incorrect predictions approach 0.

#### Training details and hyperparameters

Training was conducted across two Nvidia Tesla V100 GPUs for three days. We used AdamW with a learning rate of 10^−5^, gradient clipping at 0.1, a PPO clipping parameter of 0.2, and a KL penalty coefficient of 0.005. Each batch consisted of eight prompts with four sampled responses per prompt.

## 4 Results

### 4.1 Baseline LLM Performance

Due to limited resources at participating clinics, detailed free-text clinical rationales were not collected for this dataset. Instead, an optional field allowed clinicians to record their top suspected diagnosis, which was used to estimate a human baseline accuracy of 54.8% on the test set. This value likely underestimates clinician reasoning ability, as it reflects single-label guesses recorded at triage rather than full diagnostic workups. At the same time, this moderate human baseline highlights the inherent difficulty of this task, underscoring the potential for machine learning models to support clinical decision-making during triage.

The random forest (RF) and XGBoost [25] baselines achieved average accuracies of 79% and 81%, with corresponding AUCs of 0.84 and 0.85, respectively (Figure 3). These results indicate that, within our dataset, a relatively simple, structured model is able to capture meaningful signals distinguishing *viral* from *non-viral*. The most important features identified by the RF model corresponded to clinically plausible variables, with vital signs emerging as the strongest predictors (Figure S2), underscoring that its predictive performance was driven by meaningful signals rather than spurious correlations. As such, the RF model serves as a strong benchmark against which we can evaluate the capacity of large language models (LLMs) in this setting.

**Figure 3:**
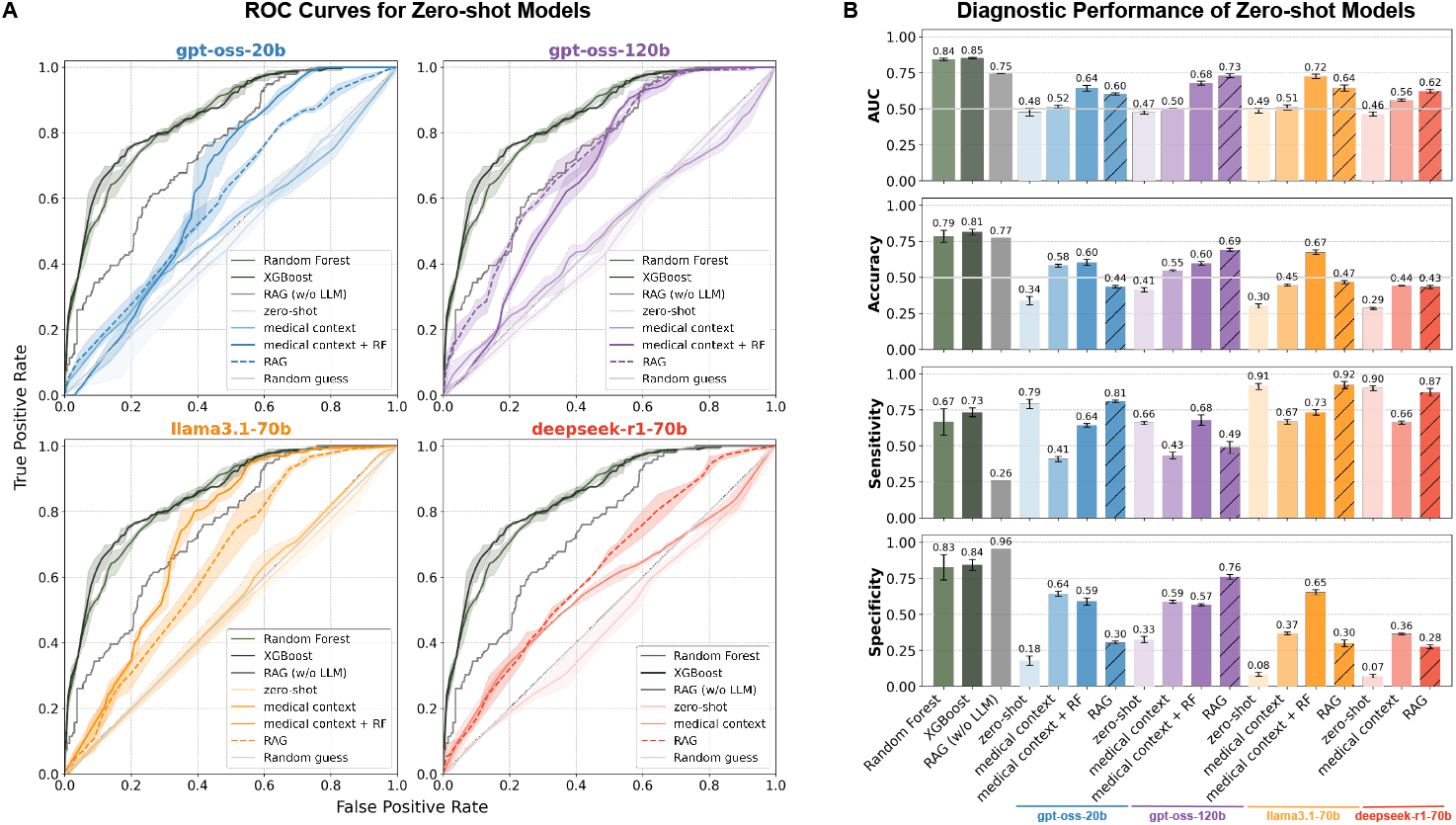
Zero-shot LLM performance. **(A)** Receiver Operating Characteristic (ROC) curves for four LLMs (`gpt-oss-20b, gpt-oss-120b, llama3.1-70b, and deepseek-r1-70b`) evaluated under four context settings: (i) applied out-of-the-box to patient data to predict *viral* vs. *non-viral* (*zero-shot*); (ii) prompted with an LLM-generated summary of 20 viral and 20 non-viral cases (*medical context*); (iii) provided with random forest (RF) predictions incorporated into the summary and patient data (*medical context + RF*); and (iv) augmented with a RAG system supplying the confirmed outcomes of the 20 nearest patients in the training set (*RAG*). Performance is benchmarked against RF, RAG (without LLM), and random guess baselines. Lines denote the mean ROC, and shaded regions indicate the minimum–maximum range across three separate inference runs for the LLMs (temperature = 0.5) and three random seeds for RF and XGBoost. The bottom-right panel reports the average area under the curve (AUC) across all methods (error bars denote standard deviation). **(B)** Overall predictive performance of each model and context setting measured as accuracy, sensitivity, and specificity. Bars indicate mean values with error bars showing the standard deviation across three separate inference runs for the LLMs (temperature = 0.5) and three random seeds for the RF and XGBoost models.

We compared the predictive performance of the baseline RF model with four SOTA open-source LLMs, gpt-oss-20b [29], gpt-oss-120b [29], llama3.1-70b [30], and deepseek-r1-70b [31], across four context settings:

1. **Zero-shot**: Models received only the patient metadata, without any additional context beyond the system prompt.
2. **Medical context**: Models were additionally provided with a distilled knowledge summary derived from 20 viral and 20 non-viral randomly chosen cases from the training data.
3. **Medical context + RF**: Models were provided with both the distilled knowledge summary and predictions from the RF model. The knowledge summary in this setting also included guidance on how to interpret RF predictions, along with the most important features identified by the RF model.
4. **Retrieval-Augmented Generation (RAG)**: Models were augmented with ground-truth labels from the 20 most similar patients to the index case, retrieved from a vector store constructed from the entire training data set.

In each setting, models were presented with a patient record and tasked with predicting *viral* vs. *non-viral* status and providing a probability of viral infection. Reported viral probabilities largely corresponded with binary yes/no predictions (Figure S3), and relative model accuracy was stable regardless of whether probability thresholds or categorical labels were used for model evaluation (Figure 3). Although the models were instructed to respond with *unknown* when insufficient data were available, they rarely did so (Figure S4).

In the baseline zero-shot condition, all LLMs performed poorly, with prediction accuracies not exceeding random guessing. Providing additional medical context via an LLM-generated summary of exemplar cases improved discrimination, and further gains were observed when either RF predictions (*medical context + RF*) or retrieval-augmented generation (*RAG*) were incorporated into the prompt (Figure 3).

Despite these gains, the RF model consistently outperformed all LLM settings (Figure 3). Importantly, when the LLMs were given direct access to RF predictions, their outputs were less accurate than the RF itself, reflecting instances where the model “overrode” the RF prediction with its own judgment. A similar pattern emerged with RAG: the `gpt-oss-20b, llama3.1-70b, and deepseek-r1-70b` models underperformed relative to the retrieved neighbors. The gpt-oss-120b model was able to closely track RAG performance but did not surpass it (Figure 3A).

Together, these findings highlight that while contextual augmentation improves LLM predictions, the models remain inferior to a simple RF baseline in this task. Moreover, even when provided with strong external signals (RF outputs or nearest-neighbor retrievals), LLMs may incorporate additional reasoning that reduces predictive accuracy relative to using those signals directly.

### 4.2 Fine-tuned LLM Performance

Fine-tuning Gemma-4B with GRPO (Gemma-4B-RL) substantially improved predictive performance, increasing AUC from 0.42 to 0.66 and accuracy from 0.26 to 0.68. Unlike the untuned model, which degenerated into predicting the same label for nearly all examples (yielding high apparent sensitivity but no practical utility), the fine-tuned model achieved balanced performance with meaningful gains in both specificity (0.01 to 0.72) and accuracy (Figure 4).

**Figure 4:**
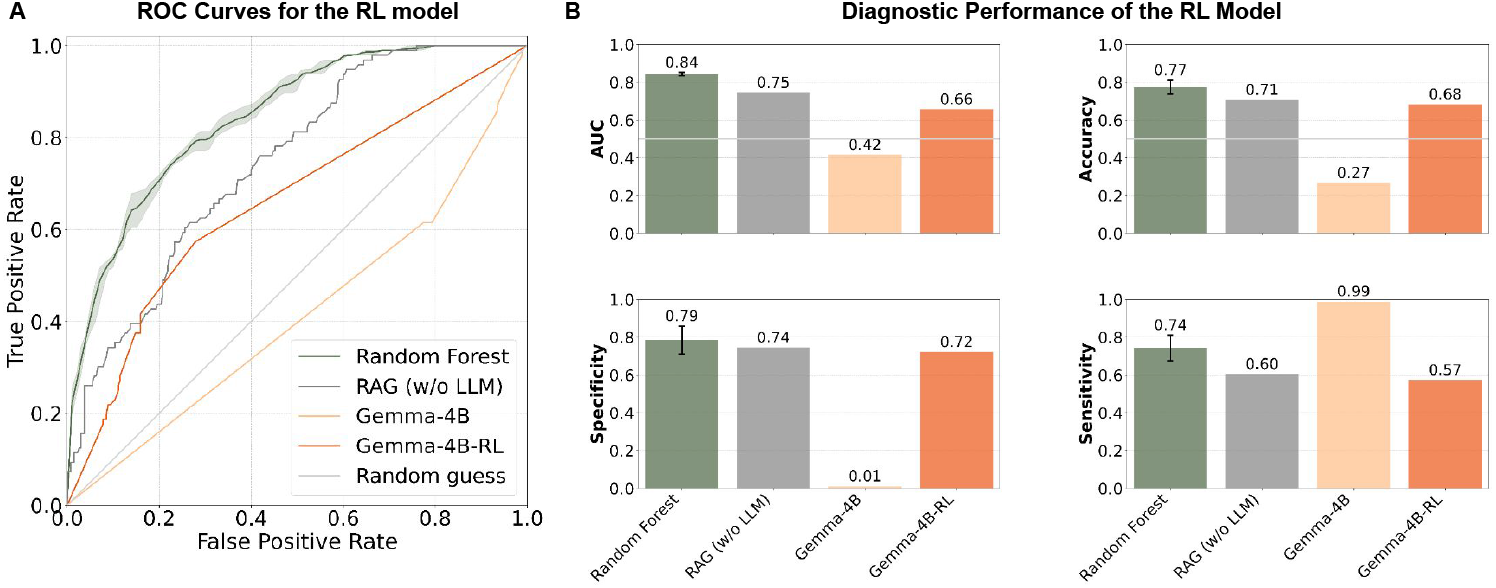
Fine-tuned LLM performance. **(A)** ROC curves showing `Gemma 4B-it` (Gemma-4B) performance versus `Gemma 4B-it` finetuned with GRPO (Gemma 4B-RL), compared against random guess, random forest (RF), and RAG (without LLM) baselines. **(B)** AUC, accuracy, specificity, and sensitivity of Gemma-4B and Gemma 4B-RL compared to the baselines. Bars indicate mean values with error bars showing the standard deviation across three random seeds for the RF model.

Relative to other open-source LLMs, Gemma-4B-RL achieves the highest AUC, with the exception of gpt-oss-120b (with medical context or RAG). However, non-LLM baselines such as Random Forest and RAG still outperform Gemma-4B-RL in both AUC and accuracy. In terms of accuracy and specificity, Gemma-4B-RL outperforms all open-source LLMs except gpt-oss-120b when combined with RAG.

We also observe that the ROC curve of Gemma-4B-RL appears close to a diagonal line, since our reward function drove the model toward making extreme predictions (near 0.05 or 0.95). This binarization amplifies discrimination at a fixed threshold but produces fewer informative gradations of probability across thresholds. This suggests that alternative reward shaping could yield smoother calibration while preserving accuracy gains. Moreover, integrating retrieval-augmented generation directly into the reinforcement learning loop represents a natural next step toward closing the gap with non-LLM baselines.

Taken together, these results highlight that targeted fine-tuning can overcome degenerate behaviors in mid-sized open-source LLMs and elevate them to competitive performance, while also pointing to clear avenues for further gains through improved reward design and hybrid RAG integration.

## 5 Conclusion

We study large language models in a setting rarely covered by existing benchmarks: **front-line, pre-diagnostic infectious disease triage**. Using first-encounter cases collected across multiple West African sites, we evaluate SOTA open-source LLMs on the operational decision of *viral* vs. *non-viral* using four context settings (medical context summaries with and without random forest predictions, and patient-RAG) alongside tabular learners. Simple tabular models recover meaningful signal from presentation-time features, whereas zero-shot LLMs underperform and remain over-confident even with RAG, which yields only modest gains. Consistent with our findings, the recently released AfriMed-QA benchmark [32] also reports that leading LLMs underperform on African medical question-answering tasks, underscoring persistent performance disparities in healthcare contexts in the global South. Aligning Gemma-4B with GRPO to a triage-oriented reward improves performance over prompting-only pipelines. To support reproducibility in low-resource settings, we focus on open models and release our evaluation harness; the dataset will be shared following de-identification and governance review. Overall, our results suggest early, low-cost clinical information can support better triage, while today’s general-purpose LLMs require targeted alignment to be useful in this pre-diagnostic context.

Our study has several limitations. (i) **Label scope:** the *non-viral* class aggregates all non-viral conditions, including bacterial, fungal, parasitic, and other diseases, into a single category, reflecting an operational decision boundary rather than a biologically distinct grouping. This is meant to reflect the on-the-ground triaging performed by clinicians. In addition, the *viral* class is not uniformly represented; common pathogens such as hepatitis B virus are overrepresented in the cohort compared to more rare viral diseases. This reflects the natural distribution of disease burden, but could introduce modeling bias. (ii) **Verification bias and noise:** not all encounters receive confirmatory testing for all pathogens. This yields high confidence in positive cases (confirmed infections), but lower confidence in negatives, where absence of confirmation may reflect under-testing rather than true absence of infection. Deep metagenomic sequencing of negative samples to confirm the absence of viral infections is ongoing. (iii) **Coverage and shift:** data comes from a limited set of sites and time periods, so broader generalization remains to be established; however, the general approach described here remains extensible to new contexts, and the value of locale-specific infectious disease models vs. global models remains to be established. (iv) **LLM formatting choices:** converting structured features to text and specific embedding/RAG settings may disadvantage LLMs; we do not exhaustively tune prompting or retrieval hyperparameters. (v) **Model inclusion:** we benchmark open-source systems for reproducibility and privacy reasons, as well as applicability of our findings in low-resource settings; closed models are excluded and can be added in future comparisons. (vi) **Training choices:** we emphasize GRPO-based alignment and do not include a supervised fine-tuning (SFT) baseline; this isolates the effect of reward-based optimization but leaves a standard SFT comparison for future work. (vii) **Safety/impact metrics:** we focus on decision performance; clinical impact, harm-avoidance, and deployment feasibility are not evaluated here.

## Data Availability

Aggregated features, metadata documentation, and sample count summaries are archived on Zenodo (DOI: 10.5281/zenodo.17065617). The complete dataset will be made publicly available after de-identification and governance review by Sentinel.

## 6 Data and code availability statement

Code is available at https://github.com/lauraluebbert/veira and model weights are available at https://huggingface.co/Sentinel-AI/neurips_grpo_model. Aggregated features, metadata documentation, and sample count summaries are archived on Zenodo (DOI: 10.5281/zenodo.17065617). The complete dataset will be made publicly available after de-identification and governance review by Sentinel.

## 7 Ethics statement

All research activities complied with relevant ethical regulations and institutional policies. Activities at the Institute for Genomic Health (IGH) were conducted under approval from the National Health Research Ethics Committee of Nigeria (NHREC/01/01/2007). Activities at the Broad Institute were conducted under approvals from the Harvard Longwood Campus Institutional Review Board (IRB24-0562, IRB24-0563).

## Acknowledgments and Disclosure of Funding

We would like to thank the clinical and laboratory teams at our Sentinel sites in Nigeria (Institute of Genomics and Global Health, Alex Ekwueme Federal University Teaching Hospital Abakaliki, Federal Medical Center Owo, General Hospital Ikorodu, and Irrua Specialist Teaching Hospital) as well as the many dedicated personnel from surrounding primary health care centers and affiliated clinics for their tireless efforts in patient enrollment, clinical data collection, laboratory analyses, quality control, and data curation, all of which were essential to the generation of this dataset.

We would like to thank Andrew White, Sam Rodriques, Jessica Wu, Geemi Wellawatte, Daniel Mukasa, Rohit Keshav Dilip, Daniel Park, and Bronwyn MacInnis for carefully reading the manuscript and providing thoughtful, constructive feedback. Their insights and suggestions greatly improved the clarity and quality of the work.

This work is made possible by support from Flu Lab and a cohort of generous donors through TED’s Audacious Project, including the ELMA Foundation, MacKenzie Scott, the Skoll Foundation, and Open Philanthropy.

L.L. is supported by funding from the FutureHouse AI-for-Science Postdoctoral Fellowship and the Eric and Wendy Schmidt Center at the Broad Institute of MIT and Harvard. Work at FutureHouse and at the Eric and Wendy Schmidt Center is supported by the generosity of Eric and Wendy Schmidt.

Y.E. is also supported by funding from the Eric and Wendy Schmidt Center at the Broad Institute of MIT and Harvard.

A.S.R is supported in part by award T32GM144273 from the National Institute of General Medical Sciences. The content is solely the responsibility of the authors and does not necessarily represent the official views of the National Institute of General Medical Sciences or the National Institutes of Health.

P.C.S. holds several patents related to diagnostic technologies and is a cofounder and equity holder in Delve Biosciences and Lyra Labs, a board member and equity holder in Polaris Genomics, and an equity holder of NextGenJane. P.C.S was formerly a co-founder of Sherlock Biosciences and board member of Danaher Corporation, until December 2024. All potential conflicts are managed in accordance with institutional policy.

The authors declare that they have no additional competing interests beyond those disclosed above.

## A Appendix

## A.1 Additional Methods

### Common-sense filters for data preprocessing

Patient age was restricted to non-negative values, systolic blood pressure (SBP) to ≤200 mmHg, diastolic blood pressure (DBP) to ≤150 mmHg, pulse rate to ≤200 beats per minute, respiratory rate to ≤60 breaths per minute, and body weight to ≤300 kg. In addition, encounter dates were required to be valid calendar entries from the year 2020 onward. Records failing these criteria were excluded (records with missing values were retained).

### Generation of training and testing splits

To generate training and testing datasets, we first excluded samples with neither a *viral* nor a *non-viral* ground truth label and retained only those with confirmed positive or negative outcomes. The binary outcome labels were extracted as the dependent variable (y), while all remaining structured clinical and epidemiological features (excluding outcome labels) were used as predictors (X). We then split the data into training (80%) and testing (20%) sets using a stratified train–test split to preserve class balance, with a fixed random seed (42) to ensure reproducibility.

### Random forest model

Numerical and categorical variables were identified based on curated data labels. Numerical variables were standardized using `StandardScaler`, while categorical variables were one-hot encoded with `OneHotEncoder` (ignoring unseen categories at test time). Preprocessing steps were implemented in a `ColumnTransformer` pipeline to ensure consistent transformations between training and testing data. Irrelevant features, such as patient identifiers, were excluded.

The model was trained on the training set using `RandomForestClassifier` with 100 trees (`n_estimators`=100). Both training and evaluation were repeated across three different random seeds (1, 42, 120) to ensure reproducibility. We also trained models on randomly permuted outcome labels (*scrambled* condition), which consistently yielded performance at random baseline (Figure S2).

Feature importance scores were extracted from trained forests. To aid interpretation, we also derived reduced feature sets (top 20, 10, and 5 features by importance determined by the random forest model trained on all features and features most frequently available in hospital settings) and retrained the random forest model on each set (Figure S2).

### XGBoost model

We implemented a gradient boosting baseline using `XGBoostClassifier`. Numerical and categorical variables were standardized and one-hot encoded using the same `ColumnTransformer` pipeline to ensure identical transformations between training and testing. Irrelevant identifiers (e.g., patient identifiers) were excluded prior to model fitting.

The XGBoost model was trained on the training split with 600 estimators (`n_estimators`=600), maximum tree depth of 5 (`max_depth`=5), learning rate of 0.05, and regularization parameters `reg_lambda=1.0 and min_child_weight`=1.0. Subsampling and column sampling ratios were set to 0.8 (`subsample and colsample_bytree`) to mitigate overfitting. Models were trained with the `binary:logistic` objective and AUC evaluation metric. Early stopping was applied with a patience of 50 rounds based on validation AUC, and training was repeated across three random seeds (1, 42, 120) for reproducibility. GPU acceleration (`tree_method=“gpu_hist”`) was enabled.

To confirm that observed performance was not driven by dataset artifacts, we also trained models on randomly permuted outcome labels (*scrambled* condition), which consistently produced AUC values at random baseline.

### Medical Context Summaries

Records from 20 *viral* and 20 *non-viral* patients were randomly chosen from the training data, converted to JSON, and grouped into batches that fit within the LLM context window. Each batch was submitted to a local LLM (`gpt-oss-120b`, temperature=0.3) together with the structured field definitions. The model was instructed to extract only high-value patterns and return a concise set of rules. Summaries were returned as bullet-point lists grouped into *viral indicators* and *non-viral indicators* (plus *RF interpretation rules* when RF predictions were included). When RF predictions were included, the batch prompt moreover included a list of the most important features identified by the RF model.

In a second stage, all batch summaries were merged with another LLM call, which deduplicated overlapping items and produced a single compact knowledge base. This distilled summary was later provided to downstream models as the medical context. The exact batch and merging prompts are provided in Box A.2 and Box A.3.

**Table A1:** User prompts used in different context settings.

| Context Setting | User Prompt |
| --- | --- |
| Zero-shot | Patient data:<br>{patient_text} |
| Medical context | Use the distilled knowledge base to guide your prediction.<br>Knowledge base:<br>{knowledge_summary}<br>Now, analyze the current patient:<br>{patient_text} |
| RAG | Viral diagnoses (1=pos, 0=neg) of the top {top_k} most similar patients:<br>{retrieved_block}<br>Now, analyze the current patient:<br>{patient_text} |

#### Box A.1

~~~
LLM System Prompt
You are an expert infectious disease physician and public health expert,
helping to prioritize patients for viral pathogen detection based on their clinical data.
The patient metadata will be structured as a JSON object with some of the following fields:
{field_definitions}
Use this additional information about the fields to inform your predictions.
From the metadata, decide:
1. Is the case viral? (yes/no)
2.Probability of viral (%)
Respond only in this exact format:
Viral: <yes / no>
Probability of viral: <percentage>
Base your answer only on the data. If unsure, output unknown.
~~~

#### Box A.2

~~~
LLM Medical Context Batch Prompt (with RF)
You are an expert clinical reasoning engine analyzing a dataset of labeled patient cases.
Each case includes:
-Structured patient metadata (symptoms, demographics, exposures, comorbidities)
-A ground truth diagnosis (‘viral_diagnosis’)
-Predictions from a random forest model (‘probability_of_viral_rf’ and ‘viral_rf’)
The RF model found the following features to be highly predictive. Critically assess their clinical plausibility:
{feature_importance_text}
Your task:
Distill only the most essential, high-value insights that will help another model predict viral vs. non-viral cases. Keep your summary short. Focus on what is consistently important and ignore less relevant patterns.
Specifically:
-Key symptom/exposure patterns strongly linked to viral infection.
-Key patterns strongly linked to non-viral cases.
-How RF predictions align or misalign with true outcomes.
-When to trust or override the RF outputs.
Output format:
A concise bullet-point list grouped into:
1. “Viral indicators”
2. “Non-viral indicators”
3. “RF interpretation rules”
Do not include explanations, background context, or verbose narrative. Only include the minimal set of rules and patterns essential for training the next model.
Here is the training data:
~~~

#### Box A.3

~~~
LLM Medical Context Merging Prompt (with RF)
You are an expert in infectious diseases and clinical data analysis.
Your task:
Merge the following batch summaries into one concise, unified clinical knowledge base that will help another model make accurate viral vs. non-viral predictions.
Include only:
-Consistent, high-value relationships between exposures, symptoms, and pathogens.
-Key patterns that strongly indicate viral or non-viral status.
-Notable exceptions and rare but important edge cases.
-Key takeaways from Random Forest predictions: when to trust or override them, and the most important features.
Output format:
A short bullet-point list grouped into:
1 “Viral indicators”
2 “Non-viral indicators”
3 “RF interpretation rules”
Instructions:
-Eliminate repetition from the batch summaries.
-Ignore low-value or inconsistent observations.
-Do not include background explanations or long narratives.
-Focus only on rules and patterns essential for model training.
Below are the batch summaries:
~~~

## B. Supplementary Figures

**Figure S1:**
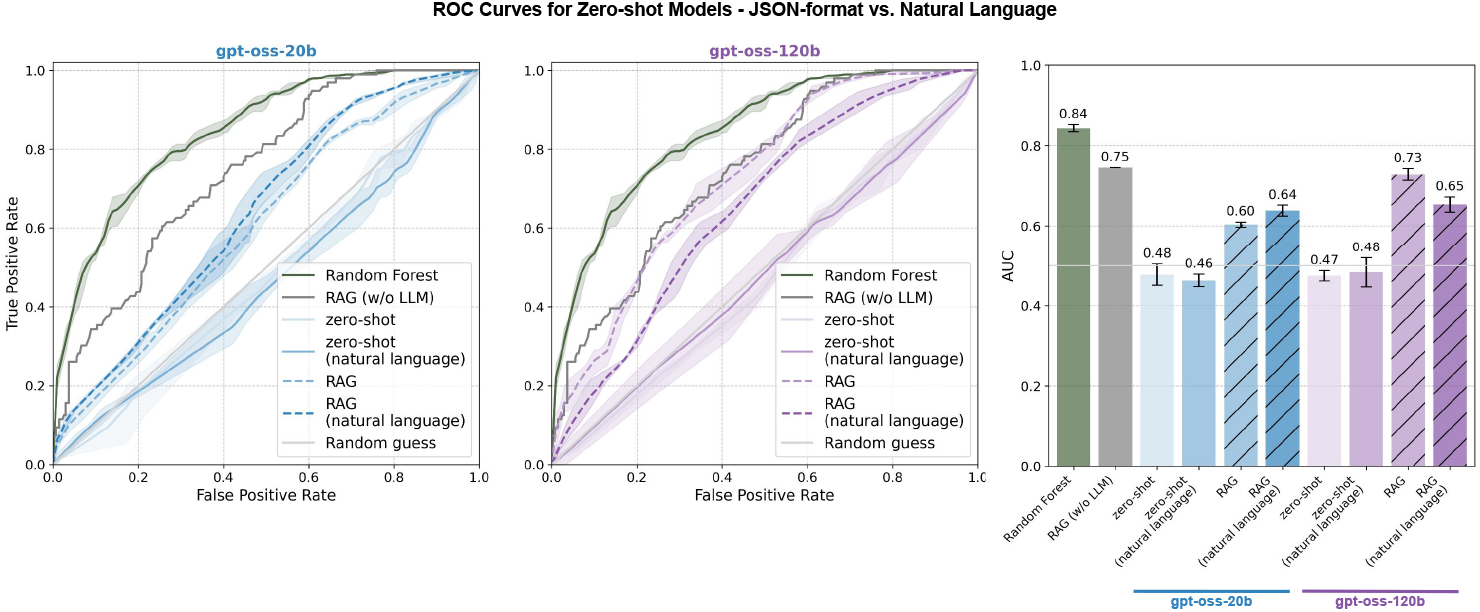
Patient data in structured JSON format vs. natural language. To rule out the possibility that model performance was limited by difficulties in interpreting patient data in structured JSON format, we also evaluated the LLMs using prompts with the patient data converted to natural language. This modification did not affect the performance of the model.

**Figure S2:**
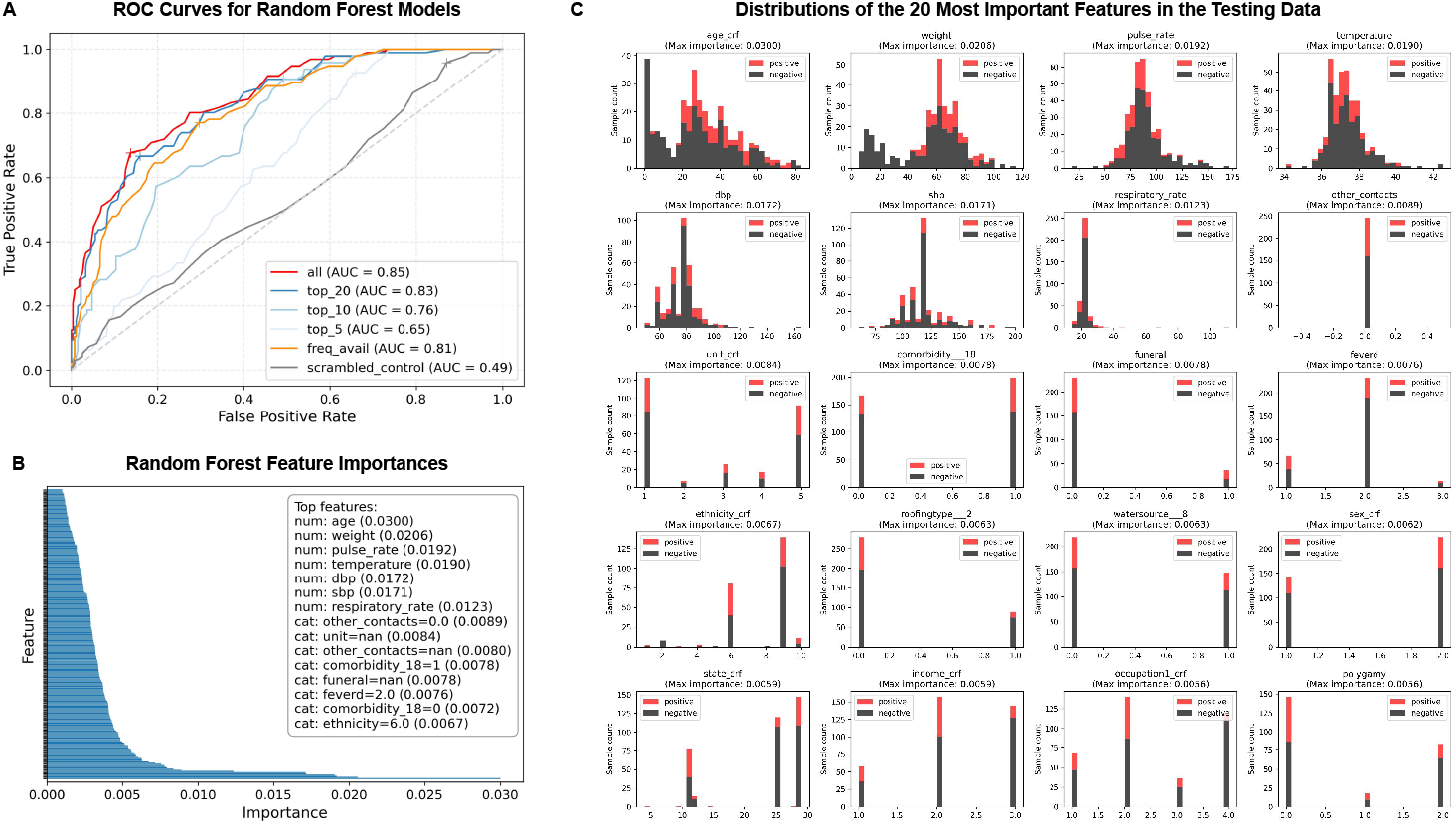
Performance and feature analysis of random forest models. **(A)** Receiver operating characteristic (ROC) curves for RF models trained on different subsets of clinical features: all features, the top 20, top 10, and top 5 as identified by the full RF model, as well as routinely collected clinical features (*freq_avail*). **(B)** Ranked feature importances of the RF model trained on all features, highlighting the strongest numerical (*num*) and categorical (*cat*) predictors. **(C)** Distributions of the 20 most important features identified by the RF model trained on all features in the testing data, stratified by ground-truth infection status: *viral* (positive) and *non-viral* (negative).

**Figure S3:**
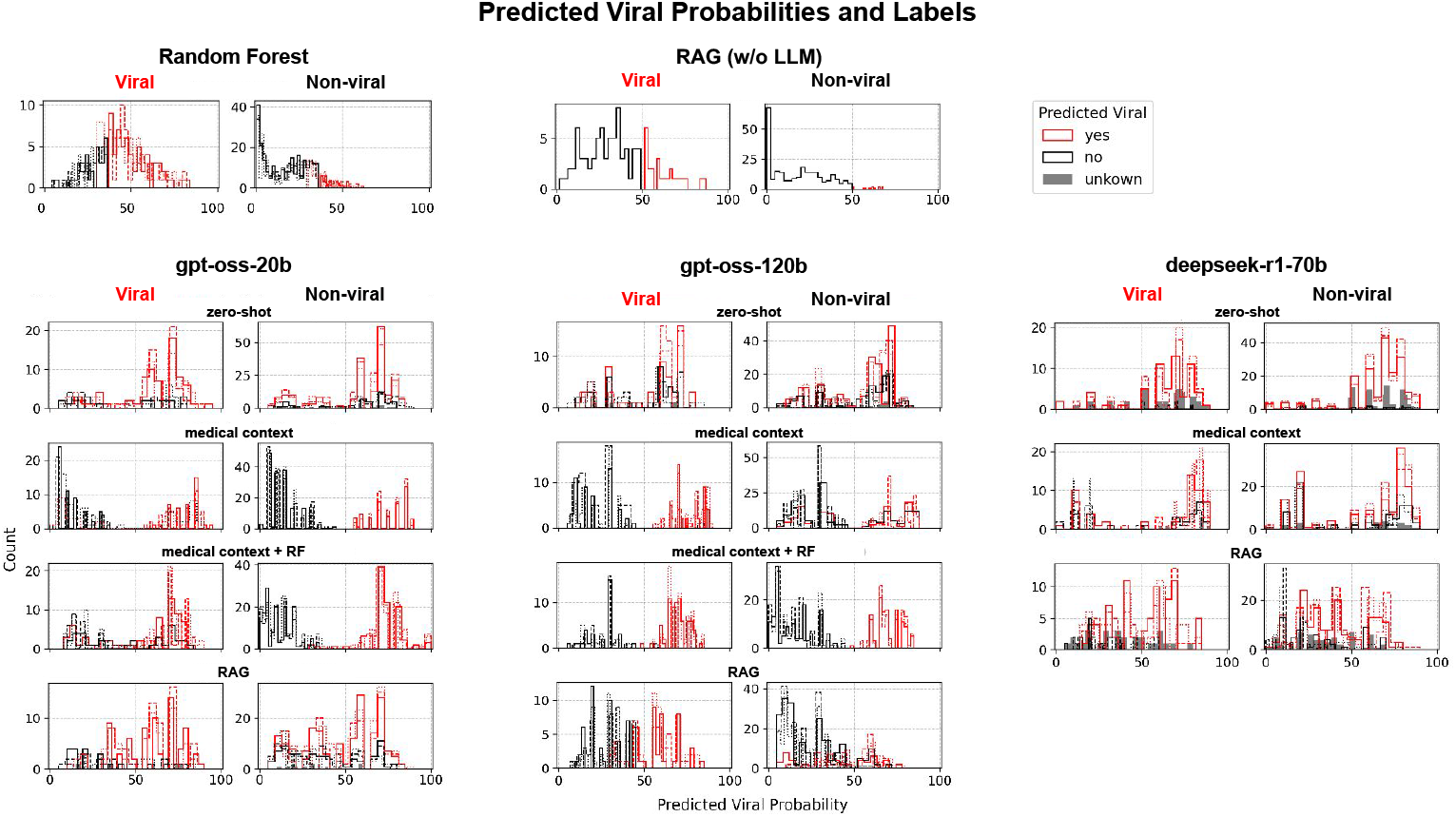
Distribution of predicted viral probabilities and binary labels across models and context settings. Each panel shows histograms of predicted probabilities stratified by ground-truth status: *viral* (left column) and *non-viral* (right column). Rows correspond to different models and context settings. The different line styles (solid, dashed, and dash–dot) correspond to three independent LLM inference runs (temperature = 0.5) or random seeds for the RF model.

**Figure S4:**
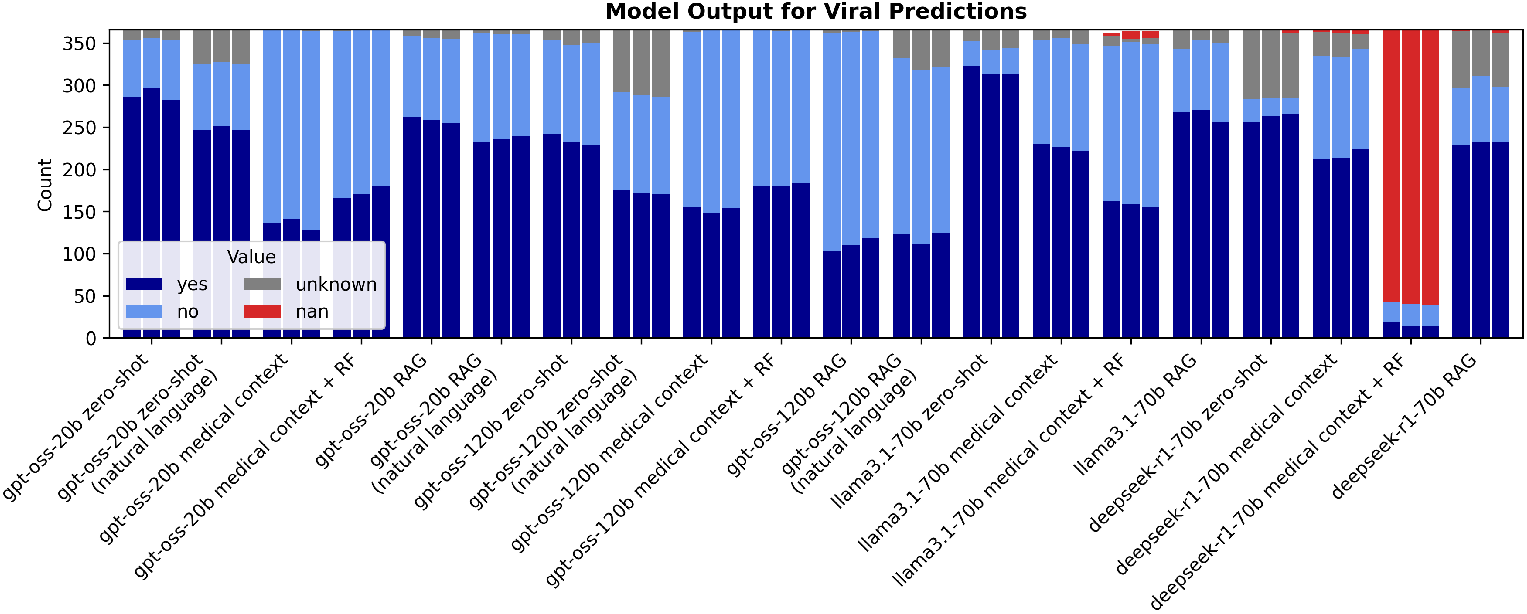
Model outputs for viral predictions across different LLMs and context settings. Bars indicate the occurrence of predicted labels (“yes”, “no”, “unknown”, or missing (“nan”)) for each model–context combination. While models were explicitly prompted to return “unknown” when insufficient data were available, this response was infrequent, with most outputs falling into binary yes/no categories. The *deepseek-r1-70b medical context + RF* configuration produced large fractions of missing values due to context window limitations deriving from the computational resources that we had available for this study, and this configuration was therefore omitted from the rest of this study.

**Figure S5:**
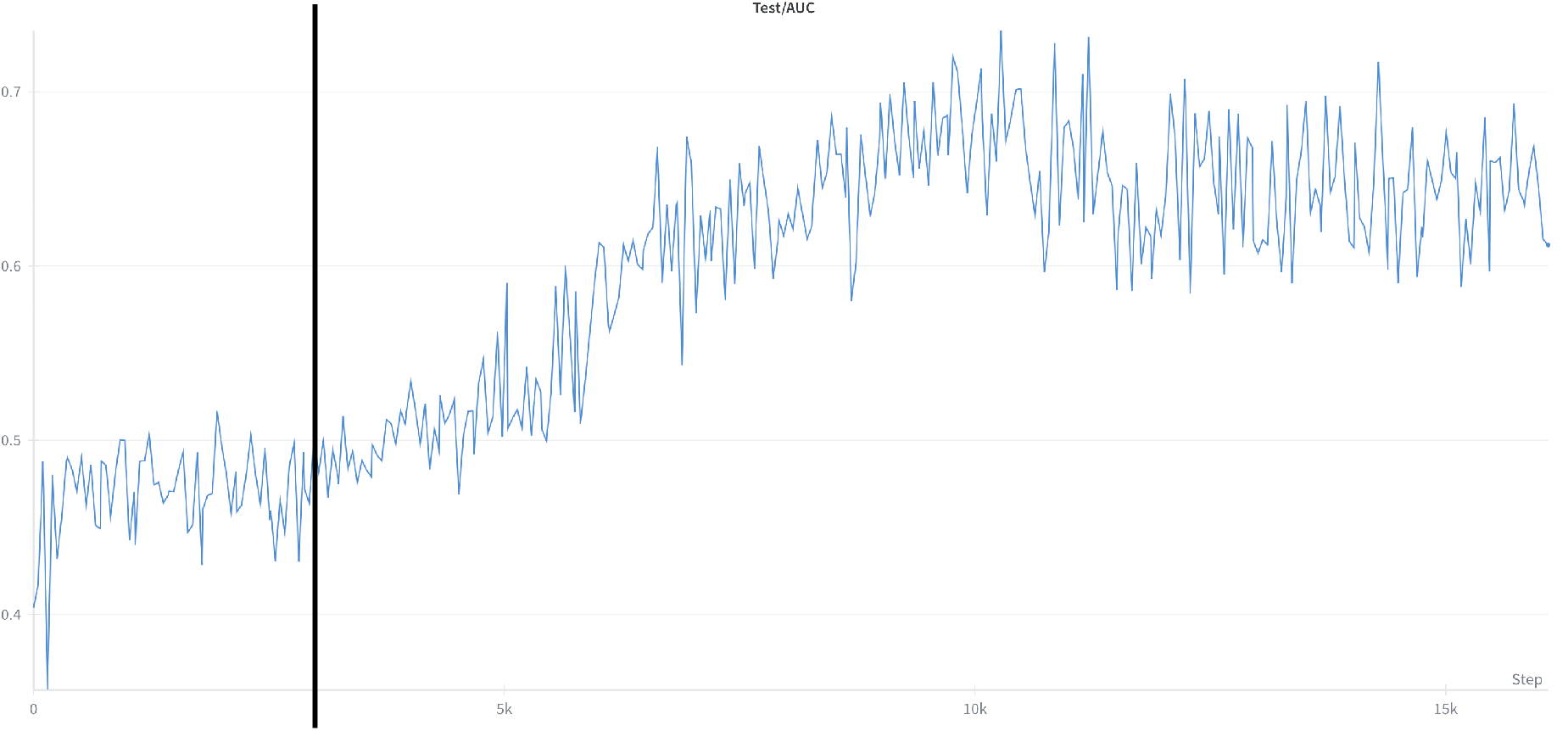
Training dynamics of Gemma-4B-RL. The model exhibits an extended plateau phase (black line) before entering a period of rapid improvement. Shown is the AUC on a balanced test set.

